# A single-center retrospective cohort study of Covid-19 medications: Remdesivir, Favipiravir, Methylprednisolone, Dexamethasone, and Interferon β1a and their combinations

**DOI:** 10.1101/2021.03.05.21251351

**Authors:** Sahand Tehrani Fateh, Sepand Tehrani Fateh, Esmaeil Salehi, Nima Rezai, Nazanin Haririan, Abdollah Asgari, Amir Salehi-Najafabadi

**Affiliations:** School of Medicine, Tehran University of Medical Sciences, Tehran, Iran; School of Medicine, Shahid Beheshti University of Medical Sciences, Tehran, Iran; M. Montazeri Hostpital, Isfahan University of Medical Sciences, Isfahan, Iran; Research Center for Immunodeficiencies, Children’s Medical Center, Tehran University of Medical Sciences, Tehran, Iran; Network of Immunity in Infection, Malignancy and Autoimmunity (NIIMA), Universal Scientific Education and Research Network (USERN), Tehran, Iran; Science and Research Branch, Islamic Azad University, Tehran, Iran; Department of Microbiology, School of Biology, University College of Science, University of Tehran, Tehran, Iran

## Abstract

Many drugs have been suggested to be used for Covid-19. A suitable and efficient choice of drug would make the course of Covid-19 easier. we have investigated the efficacy of different treatment regimen in reducing hospitalization period (HP) and mortality of 324 confirmed Covid-19 patients. Received drugs included single therapy or combinations of Methylprednisolone, Remdesivir, Favipiravir, Interferon β1a, and Dexamethasone. HP and mortality were compared between different treatment groups to evaluate efficacy of each drug. HP and mortality were also calculated for patients in each treatment group based on their underlying diseases and age. we suggest that using IFN-β1a, RDV and corticosteroids might not have a significant effect on the HP or mortality of the Covid-19 patients as it was thought before.

## Introduction

Coronavirus disease 2019 (Covid-19), caused by severe acute respiratory syndrome coronavirus 2 (SARS-CoV-2), has affected millions of individuals worldwide, leading to more than a million death since December 2019. ^1^. As SARS-CoV-19 spreads violently and takes lives, there are many efforts to develop new drugs or demonstrate the efficacy of previously developed drugs for decreasing the morbidity and mortality. Many trials, cohort studies, and meta-analyses are conducted in order to prove the efficacy of various drugs including, hydroxychloroquine, methylprednisolone (MPS), Dexamethasone (DXM), lopinavir-ritonavir, Favipiravir (FPV), interferon, and Remdesivir (RDV)^2–5^; however, in some cases, including corticosteroids and FPV, no consensus has been achieved on their efficacy for treatment of Covid-19^6–9^. Moreover, in spite of an FDA approval for RDV, different reports from all over the world are still being published to improve our understanding of its efficacy. Therefore, various studies and reports on single or combination therapies of Covid-19 are being published in order to facilitate the clarification of these therapies’ efficacy.

Herein, we conducted a retrospective cohort study on 324 confirmed Covid-19 patients assigned to different treatment groups randomly. Treatments included different combinations of drugs, including MPS, RDV, FPV, Interferon-β 1a (IFN-β1a), and DXM.

## Methods

### Study design and participant

We performed a retrospective cohort study on adult patients (age≥18), who were admitted to a hospital in Isfahan, Iran, from June 28, 2020 to November 4, 2020. Patients who received treatments were all informed and a consent is obtained. SARS-COV-2 infection was confirmed by standard reverse transcription polymerase chain reaction (RT-PCR) test, while chest computed tomography (CT) scan was also done; those who had either radiographic evidence of at least 30% pulmonary involvement or oxygen saturation of 90% or lesser were included in this study (A total of 324 patients). The treatment regimen for each group of patients was different, and it was related to the accessibility and availability of drugs in different periods of time. MPS, RDV, FPV, IFN-β 1a, and DXM, were the drugs that individually or in combination with each other had been used for the treatment of COVID-19 patients in this study. Patients’ medical history and demographic data were obtained at hospital admission. The treatment regimen and Hospitalization period (HP) for each patient were recorded.

### Procedures

The treatment was started once the patient was tested positive for SARS-COV-2 and had either radiographic evidence of at least 30% pulmonary involvement or oxygen saturation of 90% or lesser. Patients were assigned to different treatment groups. There was a total of 12 groups, including 1-patients who only received MPS; 2-patients who only received IFN-β 1a; 3-patients who only received RDV; 4-patients who only received FPV; 5-patients who received FPV and IFN-β 1a; 6-patients who received MPS and RDV; 7-patients who received MPS and RDV and DXM; 8-patients who received MPS and RDV and DXM and IFN-β 1a; 9-patients who received DXM and RDV; 10-patients who received MPS and IFN-β 1a; 11-patients who received DXM and IFN-β 1a; 12-patients who received MPS and IFN-β 1a and DXM. MPS (500mg), DXM (4mg/ml), and RDV (Loading dose: 200mg, Daily dose: 100mg) were injected intravenously to the patients, IFN-β 1a (Dose: 44µg per shot) was injected intramuscularly, and FPV (Dose: 200mg) was taken orally. The mean dose that is reported is the average number of tablets or shots of each drug that each patient received.

Patients either died or were discharged from the hospital after treatment. Patients were discharged from the hospital after recovery only, if they were tested negative for SARS-CoV-2 and had normal oxygen saturation and a normal chest CT scan. The treatment regimen and HP for each patient were recorded.

### Outcome

The study’s primary outcome was composed of mortality and mean HP for each treatment group, and the mean dose of each drug, sex ratio, and mean age were calculated in each group. The before-mentioned indices were also calculated for each group’s minor classifications (i.e., no underlying disease, underlying disease and its sub-groups, age groups), which can be found in the supplementary data.

### Statistical analysis

We compared the mean HP and mortality between different treatment groups to see if there is any significant difference between different drugs’ performance in reducing the mean HP or mortality. To compare the mean HP of the two groups, we first compared the mean age and also the mean dose of drugs between the two groups to avoid any confounding variables. We also excluded patients who died during the study when comparing the mean HP between groups. We only compared groups that their treatment regimen differed only by one drug, including 1-MPS/IFN-β1a/DXM group and IFN-β1a/DXM group 2-MPS group and MPS/IFN-β1a group 3-MPS/RDV/DXM group and MPS/RDV group 4-MPS/RDV/DXM group and RDV/DXM group 5-MPS group and MPS/RDV group 6-FPV group and FPV/IFN-β 1a group.

We used the Anderson-Darling test to assess the normality of each variable in each group, and since almost none of them were normally distributed, we used the Mann-Whitney U test for continuous variables to assess the differences in means of two groups, whereas we used fisher’s exact test for categorical variables. We did statistical analyses using R language (4.0.2 ed,2020) implemented in R studio (1.3.1093 ed,2020)^10,11^ and Microsoft Excel software. The significance for all statistical analyses was defined as p < 0.050.

## Results

A total of 324 patients was included in this study. The mean age, mean HP, and mortality for these patients were 61.37±16.76 and 7.22±4.23, and 27.47%, respectively. One hundred six of these patients had no underlying disease, and their mean age, mean HP, and mortality were 49.5±16.53, 7.25±4.74 and 16%, respectively. These indices were 67.14±13.54, 7.21±3.96, and 33.02%, respectively, for patients with underlying diseases of different kinds (n=218). There were 53 patients with hypertension, 41 with diabetes, 24 with hyperlipidemia and diabetes and hypertension, 20 with diabetes and hypertension, 19 with Heart disease, 16 with hyperlipidemia and hypertension, and patients with other diseases, which can be found in supplementary data. The indices, as mentioned above for each disease and also age groups, can be found in supplementary data.

**Figure 1.**
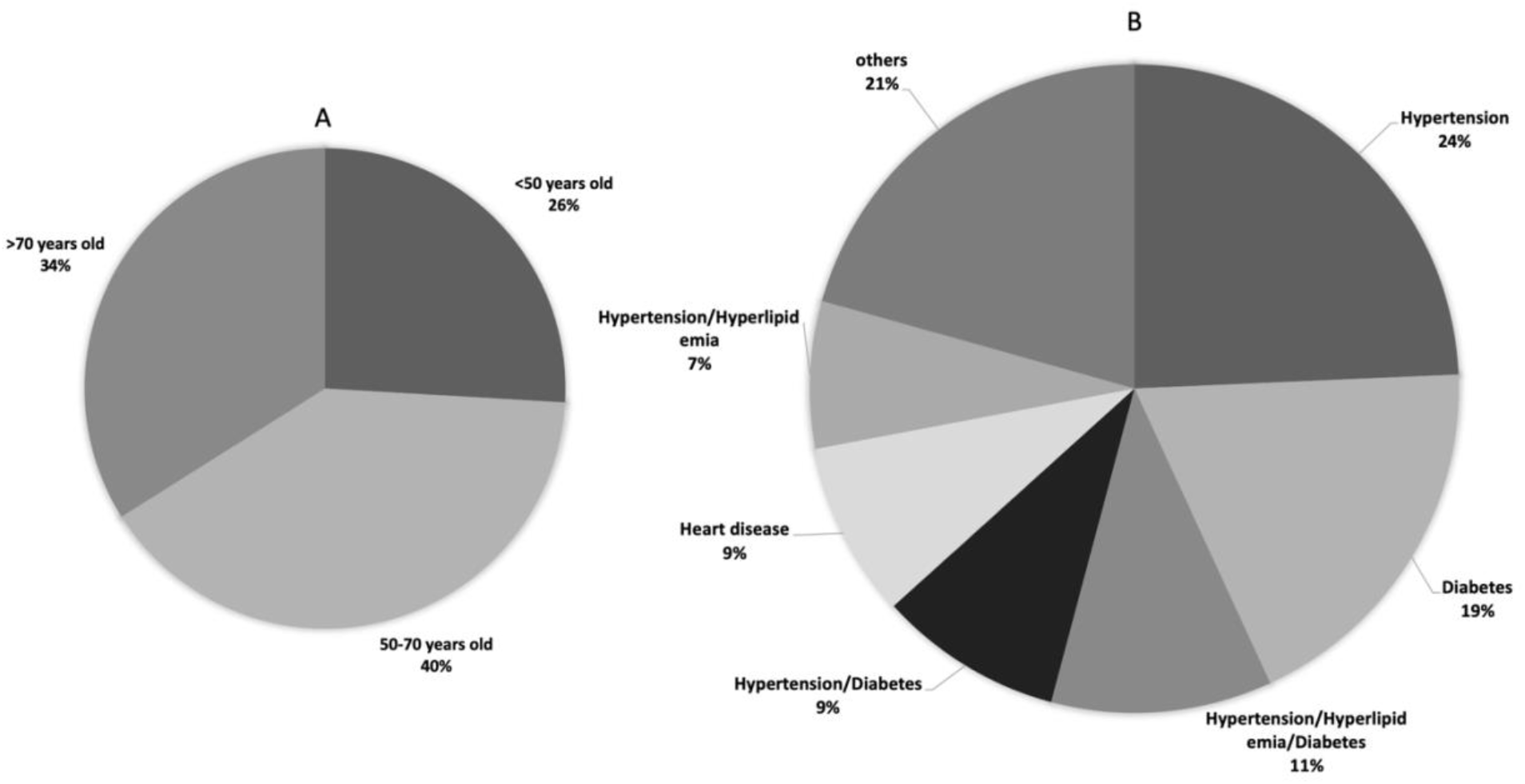
(A) Classification of 324 patients in terms of age. (B) Classification of these patients in terms of the six most common underlying diseases.

### MPS

Mean age, mean dose, and mean HP for the patients under MPS (n=77) were 63.07± 15.39, 3.71±2.86, and 7.49±4.04, respectively, and 32.46% is calculated to be the mortality. Twenty-four of these patients had no underlying diseases, and their mean age, mean dose, mean HP, and mortality were 54.67±17.14, 4.41±3.98, 7.04±3.36, and 41.66%, respectively. These indices were 66.88±12.99, 3.39±2.14, 7.69±4.33, and 45.28%, respectively, for patients with underlying diseases of different kinds (n=53). The patients with underlying diseases were also further classified into different diseases, and the mean age, mean dose, mean HP, and mortality of each sub-group can be found in the supplementary data. These patients were also classified into three age groups, including <50years old, 50-70years old, and >70 years old. Patients in the <50years old age group (n=18) had mean Age, mean dose, mean HP, and mortality of 41±5.94, 4.78±4.49, 7±3.06, and 5.56%. Patients in the 50-70 years old age group (n=29) had mean Age, mean dose, mean HP, and mortality of 61.06±5.50, 3.65±2.20, 8±4.71, and 27.58%. Patients in the >70 years old age group (n=30) had mean Age, mean dose, mean HP, and mortality of 78.26±5.09, 3.13±1.96, 7.3±3.94, and 53.34%.

### MPS and IFN-β1a

Thirty-three patients in total were taking the combination of MPS and IFN-β1a, which mean age, mean dose of MPS, mean HP, and mortality for these patients were 63.21±18.17, 3.78±3.24, 5.90±2.62, and 18.18% respectively, and all patients received one dose of IFN-β1a. Among these patients, those with no underlying diseases (n=14) had mean age, mean dose, mean HP, and mortality of 50.14±19.57, 3.35±1.82, 5.64±3.02, and 14.28%, respectively, while these indices for the patients with underlying diseases (n=19) were 72.84±8.94, 4.10±4.01, 6.10±2.35, and 21.05% respectively. The patients with underlying diseases were also further classified into different diseases, and the mean age, mean dose, mean HP, and mortality of each sub-group can be found in the supplementary data. These patients are also classified into three age groups, including <50 years old, 50-70 years old, and >70 years old. Individuals in the age group of <50 years old (n=7) had the mean age, mean dose of MPS, mean HP, and mortality of 35.14±8.41, 4.14±2.03, 7±3.31, and 14.28%. Individuals in the age group of 50-70 years old (n=10) had a mean age, mean dose of MPS, mean HP, and mortality of 58.4±8.41, 2.4±1.57, 4.7±2.54, and 20%. Individuals in the age group of >70 years old (n=16) had mean age, mean dose of MPS, mean HP, and mortality of 78.5±5.26, 4.5±4.17, 6.18±2.19, and 18.75%.

### RDV

A total of 6 patients with mean age, mean dose, mean HP of 62.5±22.33, 4.34±1.75, 10.16±5.67 were only on RDV during their course of the disease. mortality was 50% in this group. The patients with no underlying disease (n=3) mean age, mean dose, mean HP, and mortality were 42.66±7.57, 3.34±2.08, 7.33±6.11, and 33.34%, respectively, while these values for the patients with underlying diseases (n=3) were 82.33±3.05, 5.34±0.57, 13±4.35, and 66.67%.

### RDV and MPS

Mean age, mean dose of RDV, mean dose of MPS, and mean HP for the patients under the combination of RDV and MPS (n=15) were 51.8±14.67, 4.8±1.78, 6.13±5.24, and 9.6±3.69 respectively, and the mortality for these patients was 40%. Among these patients, four patients had no underlying diseases, while 11 of them had underlying diseases. The Composition of underlying diseases and values for mean age, mean dose, mean HP, and mortality of either the patients with or without underlying diseases can be found in the supplementary data.

### RDV, DXM, MPS, and IFN-β1a

A total of 7 patients took the combination of RDV, DXM, MPS, and IFN-β1a during their course of treatment. The mean dose of RDV, DXM, MPS, IFN-β1a was 5±1.41, 10±6.78, 3.71±2.62, 1.71±1.11, respectively. Mean age, mean dose, and mortality for this group were 53.71±16.10, 7.28±3.72, and 57.14%, respectively. Four of these patients had underlying diseases while the other 3 had no underlying diseases, and the mean age, mean dose, mean HP, and mortality of these two sub-groups can be found in the supplementary data.

### RDV and DXM

Thirty-two patients were taking the combination of RDV and DXM with the mean dose of 4.56±1.84 and 8.93±6.92. Mean age, mean HP, and mortality for these patients were 54.40±13.47, 6.62±3.32, and 28.12%, respectively. Sixteen patients with no underlying diseases were taking RDV and DXM with the mean dose of 4.62±1.96 and 8.93±8.11, and their mean age, mean HP, and mortality were 48.81±13.25, 6.43±3.53, and 25%, respectively. Other 16 patients with underlying diseases were taking RDV and DXM with the mean dose of 4.5±1.78 and 8.93±5.77, and their mean age, mean HP, and mortality were 60±11.52, 6.81±3.20, and 31.25%, respectively. The patients with underlying diseases were also further classified into different diseases, and the mean age, mean dose, mean HP, and mortality of each sub-group can be found in the supplementary data. These patients were also classified into three age groups, including <50 years old, 50-70 years old, and >70 years old. Patients in the <50years old age group (n=11) had the mean age, mean HP, and mortality of 39.72±7.90, 6.36±2.69, and 9.09%. Patients in the 50-70years old age group (n=19) had mean age, mean HP, and mortality of 60.10±5.79, 6.63±3.83, and 42.10%. Patients in the >70years old age group (n=2) had mean age, mean HP, and mortality of 81±0.00, 8±1.41, and 0%. The mean dose of each drug of the current combination for all age sub-groups can be found in the supplementary data.

### RDV, DXM, and MPS

Twenty-nine patients in total were taking the combination of RDV, DXM, MPS as their Covid-19 treatment with the mean dose of 5.20±2.54, 6.44±6.81, 4.96±5.06, respectively. Mean age, mean HP, and mortality for these patients were 52.55±16.10, 8.44±5.86, and 34.48%, respectively. Mean age, mean HP, and mortality for the patients with no underlying diseases (n=12) were 40.75±13.02, 11.34±7.94, and 8.34%, respectively. Mean age, mean HP, and mortality for the patients with underlying diseases (n=17) were 52.55±16.10, 8.44±5.86, and 34.48%, respectively. Mean dose of RDV, DXM, MPS for the patients with no underlying disease were 5.75±2.86, 8.58±9.37, 6.34±5.69, respectively. The mean dose of RDV, DXM, MPS for the patients with the underlying disease were 4.82±2.29, 4.94±3.86, 4.00±4.50, respectively. The patients with underlying diseases were also further classified into different diseases, and the mean age, mean dose, mean HP, and mortality of each sub-group can be found in the supplementary data. These patients were also classified into three age groups, including <50years old, 50-70years old, and >70 years old. Patients in the <50years old age group (n=12) had mean Age, mean HP, and mortality of 36.08±8.02, 11.25±7.80, and 25%. Patients in the 50-70years old age group (n=12) had mean Age, mean HP, and mortality of 60.16±5.00, 5.83±2.91, and 41.66%. Patients in the >70years old age group (n=5) had the mean Age, mean HP, and mortality of 73.8±4.08, 8±2.34, and 40%. The mean dose of each drug of the current combination for all age sub-groups can be found in the supplementary data.

### IFN-β1a

A total of 8 patients with mean age, mean dose, mean HP of 69±14.92, 2.25±1.03, 4.37±1.50 was only on IFN-β1a during their course of the disease, and the mortality was 0% in this group. Only one patient with no underlying diseases was in this group, while mean age, mean dose, mean HP, and mortality values for the patients with underlying diseases (n=7) were 70.85±15.09, 2.14±1.06, 4.57±1.51, and 0%.

### IFN-β1a and DXM

Thirty-seven patients were taking the combination of IFN-β1a and DXM with the mean dose of 2.02±1.46 and 6.67±7.76, respectively. Mean age, mean HP, and mortality for these patients were 68.89±16.72, 5.43±3.79, and 16.21%, respectively. Mean age, mean HP, and mortality for the patients with no underlying diseases (n=4) were 51.25±6.60, 4±2.82, and 0%, respectively. Mean age, mean HP, and mortality for the patients with underlying diseases (n=33) were 71.03±16.33, 5.60±3.89, and 18.18%, respectively. The mean dose of IFN-β1a and DXM for the patients with no underlying disease was 2.5±1.73 and 6.5±5.80, respectively. The mean dose of IFN-β1a and DXM for the patients with the underlying disease was 1.96±1.44 and 6.69±8.04, respectively. The patients with underlying diseases were also further classified into different diseases, and the mean age, mean dose, mean HP, and mortality of each sub-group can be found in the supplementary data. These patients were also classified into three age groups, including <50 years old, 50-70 years old, and >70 years old. Patients in the <50 years old age group (n=5) had a mean age, mean HP, and mortality of 37.6±9.20, 5.4±1.81, and 0%. Patients in the 50-70 years old age group (n=13) had mean age, mean HP, and mortality of 62±3.51, 4.61±3.37, and 7.6%. Patients in the >70 years old age group (n=19) had mean age, mean HP, and mortality of 81.84±6.99, 6±4.43, and 26.31%. The mean dose of each drug of the current combination for all age sub-groups can be found in the supplementary data.

### IFN-β1a, DXM, and MPS

Fifty patients in total were taking the combination of IFN-β1a, DXM, and MPS with the mean dose of 2.86±1.59, 7±6.23, 3.5±3.38, respectively. Mean age, mean HP, and mortality for these patients were 66.52±15.72, 7.4±4.11, and 32%, respectively. Mean age, mean HP, and mortality for the patients with no underlying diseases (n=15) were 56.2±18.73, 8.4±6.08, and 33.34%, respectively. Mean age, mean HP, and mortality for the patients with underlying diseases (n=35) were 70.94±12.02, 6.97±2.91, and 31.42%, respectively. The mean dose of IFN-β1a, DXM, and MPS for the patients with no underlying disease (n=15) were 2.8±2.17, 6.93±7.01, and 4.13±5.08, respectively. The mean dose of IFN-β1a, DXM, and MPS for the patients with the underlying disease was 2.88±1.30, 7.02±5.98, and 3.22±2.36, respectively. The patients with underlying diseases were also further classified into different diseases, and the mean age, mean dose, mean HP, and mortality of each sub-group can be found in the supplementary data. These patients were also classified into three age groups, including <50 years old, 50-70 years old, and >70 years old. Patients in the <50 years old age group (n=7) had a mean age, mean HP, and mortality of 38.14±8.05, 8.71±7.80, and 14.28%. Patients in the 50-70years old age group (n=20) had mean age, mean HP, and mortality of 61.05±5.76, 6.85±3.13, and 30%. Patients in the >70years old age group (n=23) had mean age, mean HP, and mortality of 79.91±5.77, 7.47±3.42, and 39.13%. The mean dose of each drug of the current combination for all age sub-groups can be found in the supplementary data.

### FPV

Eighteen patients were taking FPV during their course of treatment, and the mean age, mean dose, mean HP, and mortality of them were 54±12.37, 25.83±16.80, 6.67±3.34, and 5.6%, respectively. Six patients had no underlying diseases and their mean Age, mean dose, mean HP, and mortality were 42.17±13.86, 24.27±11.21, 4.5±1.38, and 0%, respectively, while these indices for the other 12 patients with underlying diseases were 59.92±5.87, 26.67±19.41, 7.75±3.54, and 8%. The patients with underlying diseases were also further classified into different diseases, and the mean age, mean dose, mean HP, and mortality of each sub-group can be found in the supplementary data. These patients were also classified into three age groups, including <50 years old, 50-70 years old, and >70 years old. Patients in the <50 years old age group (n=6) had the mean age, mean dose, mean HP, and mortality of 39.33±8.85, 27.83±13.03, 4.5±1.38, and 17%. Patients in the 50-70years old age group (n=12) had mean age, mean dose, mean HP, and mortality of 61.33±4.98, 24.83±18.86, 7.75±3.54, and 0%. No patients in this medication group were under 70 years old.

### FPV and IFN-β1a

Twelve patients were under the combination of FPV and Interferon β1a and mean age, mean dose of FPV, mean HP, and mortality of them were 62.42±21.35, 22.5±14.39, 10.75±6.15, and 25%, respectively, and all patients received one dose of IFN-β1a. Patients with no underlying diseases (n=4) had mean age, mean dose, mean HP, and mortality of 46±23.71, 22.25±17.73, 6.75±2.06, and 0% respectively, while these indices for the patients with underlying diseases (n=8) were 70.62±15.63, 22.62±13.81, 12.75±6.63, and 37.5%. The patients with underlying diseases were also further classified into different diseases, and the mean age, mean dose, mean HP, and mortality of each sub-group can be found in the supplementary data. These patients were also classified into three age groups, including <50 years old, 50-70 years old, and >70 years old. Patients in the <50 years old age group (n=4) had mean age, mean dose of FPV, mean HP, and mortality of 38.50±14.20, 23.50±7.68, 6.25±2.22, and 25%. Only two patients were in the 50-70 years old age group (n=2) with mean age, mean dose of FPV, mean HP, and mortality of 58±9.90, 8.5±10.61, 10.5±4.95, and 0%. Patients in the >70 years old age group (n=6) had mean age, mean dose of FPV, mean HP, and mortality of 79.83±5.71, 26.50±17.31, 13.83±6.91, and 33%.

### Statistical analysis of differences of mean HP and mortality between treatment groups

A comparison between the mean dose of IFN-β1a and DXM and mean age of IFN-β1a/DXM medication group and IFN-β1a/DXM/MPS medication group revealed that there is no significant difference between the mean dose of DXM and mean age of both groups (p-value>0.05) while the mean dose of IFN-β1a is significantly different (p-value <0.05). Moreover, although there is a significant difference between the mean HP of both groups (P-value <0.05), the difference between the mortality of them is insignificant (P-value >0.1). By comparing the difference of mean age and mean dose of MPS in the MPS medication group and IFN-β1a /MPS medication group, it is revealed that both differences are insignificant (P-value >0.05). Moreover, the differences between the mean HP and mortality of these two groups are found to be insignificant (P-value >0.05). The same comparison was made between the MPS medication group and the RDV/MPS medication group, and the result indicated that there is no significant difference between mean age, mean dose, mean HP, and mortality of each group (P-value >0.05). Comparison between RDV/DXM medication group and RDV/DXM/MPS medication group revealed that there is no statistically significant difference between the mean age, mean HP, mean dose of RDV, mean dose of DXM, and the mortality of both groups. In addition, there was no statistically significant difference between the RDV/MPS medication group and the RDV/DXM/MPS medication group in terms of mean age, mean HP, mean dose of RDV, and MPS and mortality (P-value >0.05). The difference between mean age, mean dose of FPV, mean HP, and mortality of FPV medication group and FPV/IFN-β1a medication group were calculated, and they found to be statistically insignificant; however, the calculated P-value related to the difference of mean HP of these two groups was 0.05054.

**Figure 2.**
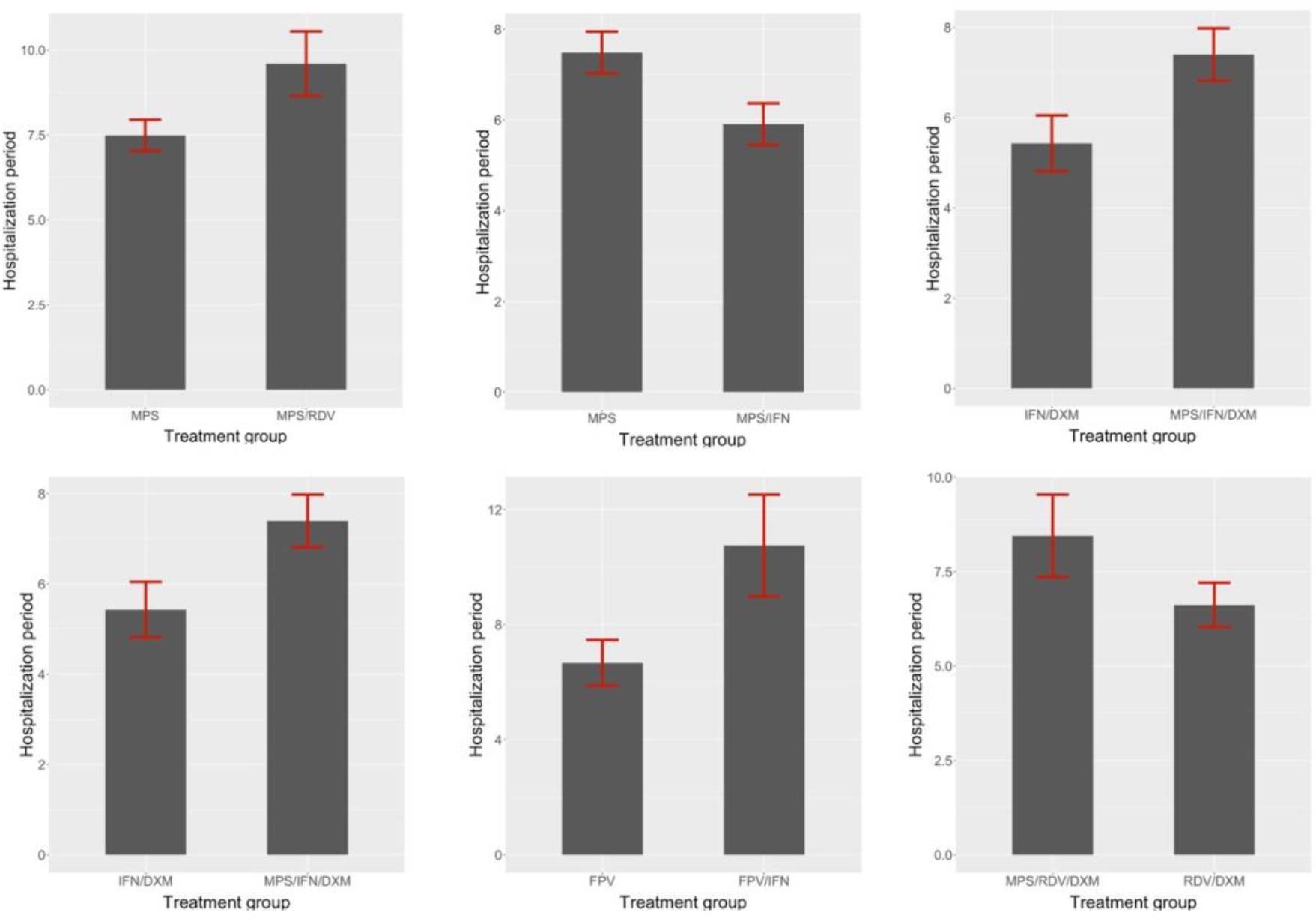
Comparison of different medication regimens in terms of their HP

**Figure 3.**
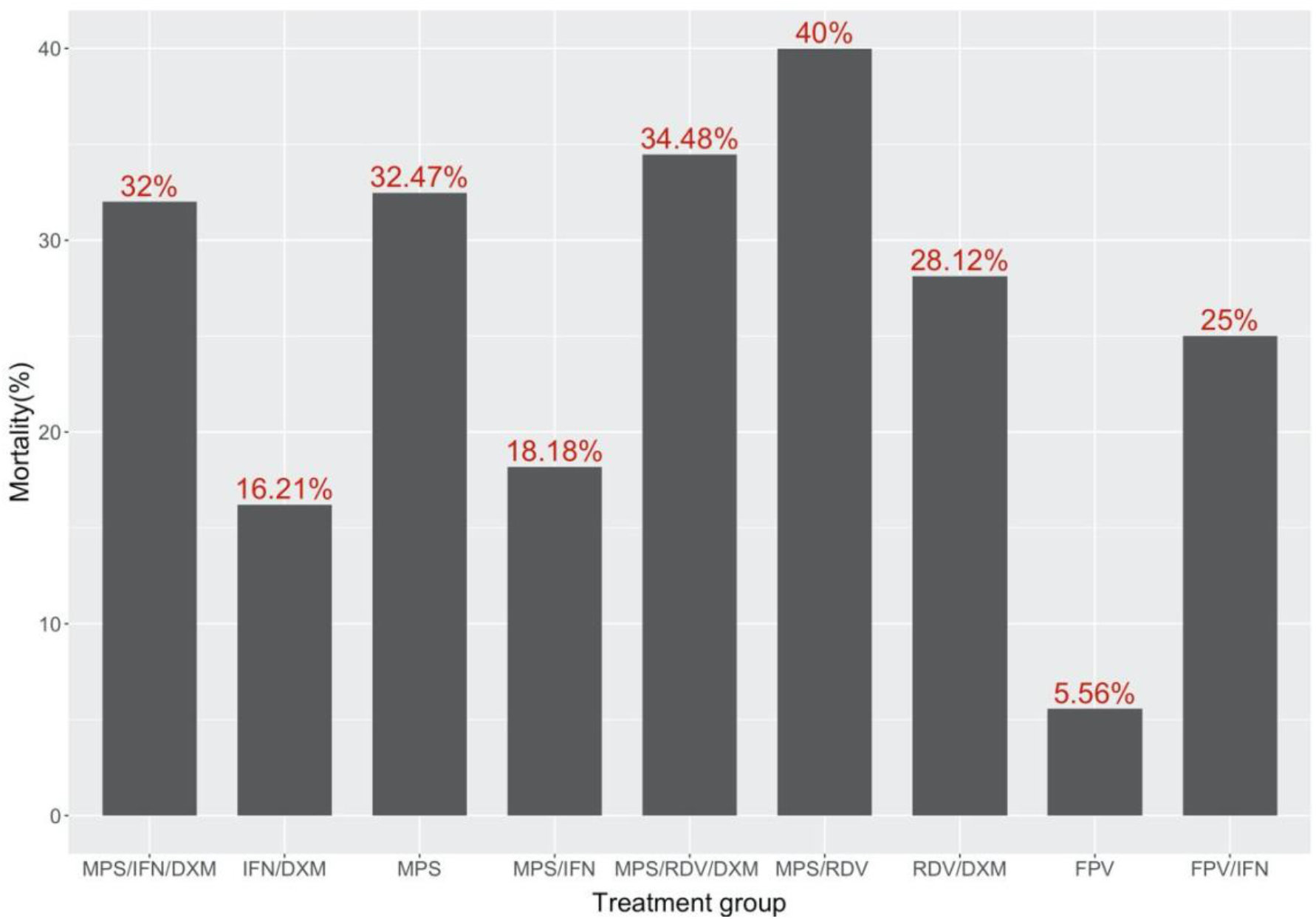
Comparison of different medication regimens in terms of the mortality of patients

## Discussion

Since the outbreak of the coronavirus, many therapeutic agents have been suggested for the treatment of COVID-19. RDV, FPV, corticosteroids, and IFN are among the most used drugs in Iran. They inhibit viral activity by different mechanisms, including inhibiting the RNA-dependent RNA polymerase, enhancing the immune system, and other unknown mechanisms. RDV, which was initially developed as a treatment for Ebola infections, is a monophosphate prodrug, and its active metabolite interferes with the action of RNA-dependent RNA polymerase leading to an inhibition of viral RNA production^12^. RDV has been approved for COVID-19 emergency treatment by the FDA on October 22, 2020^13^. The approval of RDV was supported by data from three different randomized clinical trials that included patients hospitalized with mild to severe COVID-19; however, there are some controversial data from other studies^14–17^.

FPV is another candidate for Covid-19 treatment which (T-705; 6-fluoro-3-hydroxy-2-pyrazinecarboxamide) is an antiviral agent, selectively and potently inhibits the RNA-dependent RNA polymerase (RdRp) of RNA viruses^18^. One open-labeled control study examined the effects of FPV versus Lopinavir/Ritonavir for the treatment of Covid-19. FPV group showed significant clinical outcomes, including shorter viral clearance and improvement in chest imaging. In addition, fewer adverse events were found in the FPV group than in the control group^7^. In another randomized clinical trial, patients who received FPV did not have significantly improved clinically recovery rate at day seven compared to patients who received Arbidol, but it led to significantly shorter latencies to relief for fever and cough^6^. In a study conducted by Yan *et al*., it was found that FPV antiviral activity is related to its concentration and its antiviral activity was not significant up to 100 μM in vitro, so they could not prove a benefit of the addition of FPV under the trial dosage to the existing standard treatment. Insufficient concentration of this drug can be the reason for the lack of Antiviral effects and medical benefits^19^. In conclusion, the outcomes of studies on FPV are not consistent, and further studies are needed to demonstrate its efficacy for the treatment of COVID-19.

Interferons are shown to be effective on many human coronaviruses, including MERS and SARS; however, their performance against SARS-COV-2 is not clear^20^. Therefore, many studies are conducted in order to evaluate its efficacy in Covid-19 treatment. IFN-β 1a inhibits SARS-COV-2 activity in vitro when administered after virus infection. The antiviral activity of IFN-β 1a in vitro has been shown to effectively inhibit infectious virus particles and viral RNA on treated cells compared to virus-positive infection control^21^. A randomized control trial assessed the use of IFN-β 1a in the treatment of severe COVID-19. Ninety-two patients went under randomization; Time to the clinical response was not significantly different between the IFN-taking group and control groups; however, 28-day overall mortality was significantly lower in the patients taking IFN than the control group, and also discharge rate was significantly higher in the IFN group^22^. Two other studies also demonstrated the benefit of adding IFN-β 1a to the antiviral treatment regimen in Covid-19 patients^23,24^.

Corticosteroids are also evaluated for being effective in improving the clinical course of Covid-19; however, its usage in the treatment regimen of Covid-19 is still controversial, and different publications declare differently about their benefits. Although some randomized clinical trials and meta-analysis reports have concluded that corticosteroids may cause positive effects in the clinical course of Covid-19^8,25–30^, there exist some other reports, suggesting corticosteroids are inefficient or even cause adverse effects^9,31–35^. Interestingly, even late administration of MPS pulses during the second week of the disease is also suggested to improve patients’ prognosis with Covid-19 pneumonia with features of inflammatory activity and respiratory deterioration^36^. The beneficial effects of corticosteroids in improving the clinical course of Covid-19 may be due to their immunomodulatory effects as it is shown that administration of corticosteroids (e.g., MPS) either in low doses (≤ 2 mg/kg day) or high doses would inhibit IL-6 production without delaying virus clearance^37^.

Since our study lacks a control group, we were not able to determine the efficacy of each group of drugs(s); however, we could compare the groups of drugs with only one different drug in between. It is worth noting that we first checked whether both groups’ properties except the mean HP and mortality are not significantly different. According to our data, IFN-β1a/DXM/MPS medication group had significantly higher mean HP than the IFN-β1a/DXM medication group, which may be due to the possibility that the patients in the latter group had more severe disease than the former group and hence they spend more time in the hospital in spite of the similar primary clinical status at the admission. Moreover, it can be suggested that using IFN-β1a does not present a significant improvement in a mean HP and mortality considering the fact that mean HP and mortality was not significantly different between IFN-β1a/MPS medication group and MPS medication group. Similarly, the same result can be probably concluded from comparing the FPV medication group and FPV/IFN-β1a medication group. A comparison between the MPS medication group and MPS/RDV medication group also revealed that there is no significant difference in mean HP and mortality, in addition to mean dose of MPS and mean age between these two groups in spite of the same severity of disease between patients, indicating that RDV might not have a significant effect on the clinical course of the diseases. In a similar way, by comparing other medication groups, differing in the presence of either MPS or DXM in the treatment regimen, it can be suggested that probably either MPS or DXM does not impose significant benefits on the clinical course of the disease.

After all, our study faces several limitations, including 1-Limited number of patients, 2-The existence of confounders that may not have been discovered, 3-The existence of a large variance between some parameters, 4-A slight difference in patients’ conditions in terms of severity of the disease, 5-Not considering the clinical course of the disease and the fact that some patients progress to a need for mechanical ventilation and 6-Rarely in some cases some drugs were not started from the beginning of the hospitalization.

## Conclusion

The mortality and hospitalization period for Covid-19 patients who were taking Remdesivir, Favipiravir, Methylprednisolone, Dexamethasone, and Interferon β1a or their combinations are considered and compared in this study. Many studies have been conducted on the efficacy of these drugs on clinical course and mortality of Covid-19 and no consensus has been achieved yet. This study suggests that using IFN-β1a, RDV and corticosteroids might not have a significant effect on the HP or mortality of the Covid-19 patients as it was thought before. More specific and accurate studies are needed for evaluating these drugs’ efficacy.

## Data Availability

All data are available through a request from the corresponding author.

